# COVID-19 in Bangladesh: Measuring differences in individual precautionary behaviors among young adults

**DOI:** 10.1101/2020.05.21.20108704

**Authors:** Asif Imtiaz, Noor Muhammad Khan, Md. Akram Hossain

## Abstract

**Aim:** The novel coronavirus disease 2019 (COVID-19) has already hit Bangladesh, and various control measures have been taken to flatten the epidemic curve. Due to the current demographic distribution in Bangladesh, young adults are vital to the demography of the country. Therefore, their precautionary behavior is very important to ensure the success of preventive policies. This exploratory study examined the differences in the adoption of precautionary behaviors among young adults, and estimated and compared the predictors of precautionary behavior adoption among young adults living in the capital city Dhaka and a nearby district, Tangail.

**Subject and Methods:** A total of 350 respondents from each district participated in the study. ANOVA and two-sample t-tests were utilized to detect differences in precautionary behavior across demographic groups of young adults, and quantile regression modeling was used to find the predictors of adopting precautionary behaviors and to compare these predictors between the two districts.

**Results:** Individuals who had a postgraduate education and had good mental health tended to show better precautionary behaviors in Dhaka. Female respondents from Tangail who had no psychological distress took precautionary behaviors more often than their counterparts. However, no significant differences in the adoption of precautionary behaviors to prevent COVID-19 among young adults were found between the two districts. Better self-control ability, higher education and good mental health emerged as factors that significantly shaped the precautionary behaviors of young adults in this study.

**Conclusion:** Having better knowledge did not ensure better adoption of precautionary behaviors among the participants. In effect, the government’s strong intervention to keep people at home and distant from each other and continued lockdown for several more days are probable immediate solutions. At the same time, the economic burden on lower-income people should be addressed.

## Background

Bangladesh confirmed the detection of its first three novel coronavirus disease 2019 (COVID-19)-positive cases on March 8, 2020, amid the reign of the COVID-19 terror across the globe (Paul 2020). Three days later, the World Health Organization (WHO) declared COVID-19 a pandemic because of its unrelenting and rapid spread around the world (Pogrebna and Kharmalov 2020). Bangladesh reported its first coronavirus death on March 18, 2020. The patient was a senior citizen aged almost 70 years with a history of various medical conditions (Reuters 2020). As of May 9, 2020, COVID-19 had already reached 215 countries, areas or territories affecting 3,822,382 people, among which 263,658 had lost their lives (WHO 2020). By the same date, in Bangladesh, there were 13,770 confirmed cases of COVID-19 and 214 confirmed deaths due to this pandemic (Institute of Epidemiology, Disease Control & Research (IEDCR) 2020).

This disease is currently infecting people at an exponentially increasing rate and can be transmitted by even asymptomatic or presymptomatic persons. These features have made COVID-19 harder to contain than the Middle East Respiratory Syndrome coronavirus (MERS-CoV) or Severe Acute Respiratory Syndrome (SARS-CoV) (Gates 2020). COVID-19 is highly contagious, with fever, dry cough, myalgia, fatigue, and dyspnea as its main clinical symptoms. At the severe stage of this disease, acute respiratory distress syndrome, uncontrolled metabolic acidosis, bleeding and coagulation dysfunction, and septic shock may occur (Chen et al. 2020; Zhong et al. 2020). The most vulnerable population for COVID-19 is older males with underlying diseases such as hypertension, type-2 diabetes, and cardiovascular disease (Chen et al. 2020; Huang et al. 2020). As the situation became critical and serious, the World Health Organization, in response, urged collective efforts of all countries to prevent the rapid spread of COVID-19 (WHO 2020). Moreover, the WHO decided to improve clinical and community-level knowledge regarding COVID-19 while promoting optimized use of protective equipment for infection prevention and control in community settings (WHO 2020). Lowering the predisposition of individual exposure is currently the only way to prevent COVID-19 since there is no vaccine or effective curative method to prevent this pandemic (Pogrebna and Kharmalov 2020). The approaches to reducing its transmission are mostly behavioral: handwashing; social distancing; coughing and sneezing etiquette (Haushofer and Metcalf 2020); and avoidance of touching the eyes, mouth, and nose. Several countries, including Bangladesh, declared a shutdown/lockdown to prevent the spread of COVID-19. Bangladesh deployed armed forces to ensure that people were maintaining social distancing and quarantine (ABC 2020; Daily Star 2020).

People’s adherence to these control measures is central to their success, and this adherence heavily relies upon their knowledge, attitudes, and practices toward COVID-19. Moreover, less knowledge and poor attitudes toward contagious diseases can trigger panic emotion, in effect, making matters worse while attempting to prevent the disease from spreading (Ajilore et al. 2017; Person et al. 2004; Tachfouti et al. 2012; Tao 2003). In a study conducted very recently, it was found that Chinese women of relatively high socioeconomic class possessed good knowledge, optimistic attitudes, and performed proper COVID-19 prevention practices during the time when the disease was rising rapidly (Zhong et al. 2020). It is understandable that the people of China adopted intensive precautionary behaviors to prevent COVID-19 (Li et al. 2020), and knowledge is a significant predictor of precautionary behavior adoption in general (Almutairi et al. 2015). Almost all the participants of this prior study were confident that COVID-19 would be finally controlled, and China would win against the virus. This confidence and positive attitude among the Chinese are nothing new, since the same pattern was exhibited during the SARS epidemic (Chen et al. 2003; Liu et al. 2004; Zhong et al. 2020; Zhou et al. 2004). Knowledge and perception of SARS were significantly associated with precautionary behavior adoption (Vartti et al. 2009), but the role of the public’s precautionary behaviors in controlling epidemics such as SARS requires further investigation (Bell 2004).

Persons with better self-control possess more positive coping strategies (Li et al. 2016) and sustain more satisfaction in essential life domains (Duckworth and Seligman 2005; Moffitt et al. 2011). A high level of self-control has a greater positive association with healthy and virtuous outcomes, and effective adoption of healthy behavior requires volitional control or willpower (Hoffman et al. 2008). During the days of the COVID-19 pandemic, healthy behaviors mainly include frequent hand-washing, maintaining cough and sneeze etiquette, keeping social distance and other steps required to stop its spread. Handwashing culture has been proven to be a vital predictor of the COVID-19 outbreak magnitude in different countries. Locations where citizens do not have the habit of washing their hands tend to be more exposed soon after the introduction of this virus (Pogrebna and Kharlamov 2020).

Demographic traits can significantly explain human behavior in response to the uprising of a pandemic. Never-married graduate or undergraduate males in China showed less knowledge of COVID-19, and they went to crowded places more often and wore masks less often than others during the rapid rise of the disease in mainland China (Zhong et al. 2020). Males who were less educated and older in age showed poor knowledge, negative attitudes, and poor preparedness skills for COVID-19 prevention and care (Srichan et al. 2020). Females with a better understanding of COVID-19 were more likely to adopt preventive measures than others (Kwok et al. 2020). This result has been supported by another study in which it was found that females with higher education and more knowledge about COVID-19 displayed more precautionary behaviors than their counterparts. Additionally, people with a history of chronic physical diseases underperformed precautionary behaviors to prevent themselves from being infected (Li et al. 2020).

Infection in young adults may go undetected for a long period, which makes people less aware of the intensity of the situation (Dowd et al. 2020) and almost one-third of the Bangladeshi population is aged between 15 and 29 years (National Institute of Population Research and Training (NIPORT) 2016). The WHO has set six conditions prior to ending the lockdown. Among those, two conditions are directly related to taking proper preventive measures (Chapell 2020). As the lockdown cannot go for a long time, the practice of preventive behaviors among this population requires immense concentration, and performing proper precautionary behaviors is the only difference between those with the highest and lowest probability of getting COVID-19. The present study aimed to i) determine the differences, if any, in the adoption of precautionary behaviors among young adults living in the capital city Dhaka and a nearby district, Tangail, and ii) estimate and compare the predictors of precautionary behavior adoption among the young adults living in these two districts. Despite having a public university in its center and being only 83.5 kilometers away from Dhaka, Tangail is considerably behind Dhaka in terms of vital health indicators and education (Bangladesh Bureau of Statistics (BBS) 2019), similar to all other districts (excluding the city corporations outside of Dhaka) in Bangladesh. This allows respondents from Tangail to act as a proxy of respondents of other districts, which enables us to understand and compare the dynamics of precautionary behaviors among young adults living inside and outside of the capital city. To the best of our knowledge, this study is the first behavioral study of COVID-19 in the Bangladeshi demographic setting.

## Methods

### Design and sampling

The design of this study was cross-sectional in nature. Members of the Secondary and Intermediate Level Students’ Welfare Association (SILSWA) of Dhaka and Tangail were the sampling frames for this study. SILSWA (www.silswa.org) is one of the largest student platforms in Bangladesh, with almost a million members across the country. Students from secondary to postgraduate levels are members of this association. This attribute of the association has made it a good representation of the young adults of the country. A simple random sampling strategy was adopted, and the sampling form was a questionnaire (Nooh et al. 2020). The minimum sample size was calculated using the formula n=z^2^pq/d^2^, where n= the desired sample size, z= the standard normal deviation (usually set at 1.96, which corresponds to 95% confidence interval level), p= the proportion in the target population estimated to have a particular characteristic (here p=67% from the pilot survey), q= 1-p (proportion in the target population not having the particular characteristics) and d= the degree of accuracy required (usually set at the 0.05 level). Considering all these factors along with design effect=1, the minimum sample size required for this study was 340 for each stratum, i.e., district. A list of SILSWA members was collected before the office closures. The survey participants were selected from the list by using computer-produced random numbers. The background and aims of the study were stated on the first page of the questionnaire. Declarations of anonymity and data confidentiality were also explained. Participants 16 (Zhong et al. 2020) up to 29 years of age were included in the study given their willingness to participate. The final Dhaka and Tangail samples consisted of 350 participants each.

### Measures

#### Knowledge

Participants’ knowledge about different features of COVID-19 was measured with a 12-item instrument that was developed by combining a questionnaire used in a previous study (Zhong et al. 2020) and guidelines for clinical and community management issued by the IEDCR (2020). Participants were asked to posit whether each statement was true or false. An additional “I don’t know” option was also given in the questionnaire. Specific statements were false to avoid bias. A binary scoring system was used: “1” for a correct answer and “0” for others (incorrect/unknown). Therefore, the range of the total knowledge score was from 0 to 12, where the higher the score, the better the knowledge about COVID-19. The Cronbach’s alpha of this instrument was 0.64 for the Dhaka sample and 0.72 for the Tangail sample, indicating acceptable consistency (Hinton et al. 2014).

#### Attitudes

The 2-item instrument to measure attitudes toward COVID-19 was adopted from a previous study (Zhong et al. 2020). In the first question, participants were required to state their agreement on the final control of COVID-19. The scoring system was binary, indicating “1” for choosing the option “Agree” and “0” for other options (“Disagree”/”I don’t know”). In the next question, respondents’ confidence in winning the battle against COVID-19 was measured as a dichotomous outcome (“1” for Yes and “0” for No). The Cronbach’s alpha of this instrument was 0. 85 for both the Dhaka sample and the Tangail sample, indicating very good consistency (Hinton et al. 2014).

#### Precautionary behaviors

The precautionary behaviors of the participants were measured with 9 items developed by the authors as per the guidelines circulated by the IEDCR (2020). Participants were asked to express their frequency of performing several precautionary behaviors since the COVID-19 outbreak. Responses were collected on a five-point scale ranging from “0=Never” to “4=Very Often”. The total score ranged from 0 to 36. A higher “precautionary score” indicated a better display of precautionary behavior. Cronbach’s alpha was 0.68 for the Dhaka sample and 0.71 for the Tangail sample in this study, meaning acceptable internal consistency (Hinton et al. 2014).

#### Self-control

The 13-item Brief Self-Control Scale (Tangney et al. 2004) measured the participants’ self-control on a five-point scale. Among the items, nine were reversed items. The scoring system used for nonreversed items ranged from “1=Not at all” to “5=Very much”. The scores were reversed for reversed items. Better self-control ability was indicated by a higher score (Zhong et al. 2020). The Cronbach’s alpha of this instrument was 0.73 for the Dhaka sample and 0.74 for the Tangail sample, indicating acceptable consistency (Hinton et al. 2014).

#### Mental health

Symptoms of common mental health problems were measured with a 12-item General Health Questionnaire (GHQ-12) (Goldberg 1972) since its reliability and validity have already been proven in community settings in different cultural contexts (Lindencrona et al. 2007). In our study, we used a 0-0-1-1 scoring system that generated a score ranging from 0 to 12. A higher score indicated poorer mental health. As a safe benchmark, the mean score was used as a potential threshold (Goldberg et al. 1998). The Cronbach’s alpha of this instrument was 0.74 for both the Dhaka sample and the Tangail sample, indicating good internal consistency (Hinton et al. 2014).

#### Demographics

Information on fundamental demographic traits such as sex (male, female), educational qualification (higher secondary, graduate, and postgraduate), and history of chronic/psychological disorder (yes, no) was collected.

### Data analyses and model evaluation

#### Descriptive analyses

Descriptive analyses to explore the differences in adoption of precautionary behaviors among different stratified demographic groups in terms of age, education, sex, marital status, mental health, and history of chronic/mental disorder were performed using ANOVA and two-sample t-test, as appropriate.

#### Estimation technique

Multivariate analyses were performed using the quantile regression model. Knowledge, attitude, mental health, self-control, and demographic variables acted as independent variables. Classical regression models cannot be extended beyond central locations, and quantile regression can analyze the entire conditional distribution. This appealing property has made quantile regression more popular among economists, econometricians, and biostatisticians (Machado and Mata 2005; Wei et al. 2006). Therefore, the quantile regression model was utilized since this method provides a comprehensive representation of possible causal relationships between variables (Liu and Bottai 2009).

##### Quantile regression

The quantile regression approach can characterize the entire conditional response distribution for a given set of predictors. Quantiles are considered a particular location of the distribution. Suppose that Z is defined by CDF F(z); then, the τ^th^ quantile is the value, z_τ_ such that:

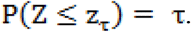

Now, the quantile function (QF) can be defined as its inverse:

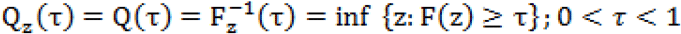

If F(.) is strictly increasing and continuous, then F^−1^(τ)is the unique real number z such that F(z) = τ (Davino et al. 2013; Gilchrist 2000). The empirical distribution function can be written as:

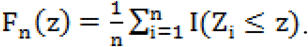

Then, the τ^th^ sample quantile is defined as:

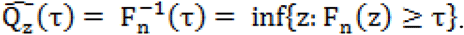

Hao and Naiman (2007) defined quantiles as specific centers of the distribution, minimizing the weighted absolute sum of deviations. Let Y be a continuous random variable with PDF f(y). Hence, the τ^th^ quantile is defined as:

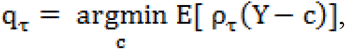

where q_τ_(.) indicates the following loss function:

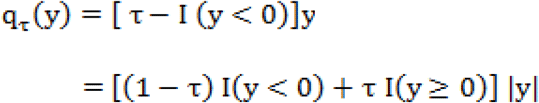

q_τ_(.) is defined as an asymmetric absolute loss function where (1 − τ) is assigned for negative deviations and τ is assigned for positive deviations.

Now, the minimization problem of the quantile function becomes:

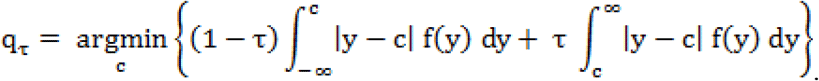

To minimize expected loss, we differentiated E [ρ_τ_(Y − c)] regarding c and then set the derivative equal to zero. This gives the solution for this minimization problem:

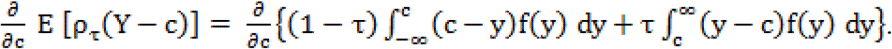

Leibniz’s rule for differentiation is applied under the integral signs in the above equation and becomes:

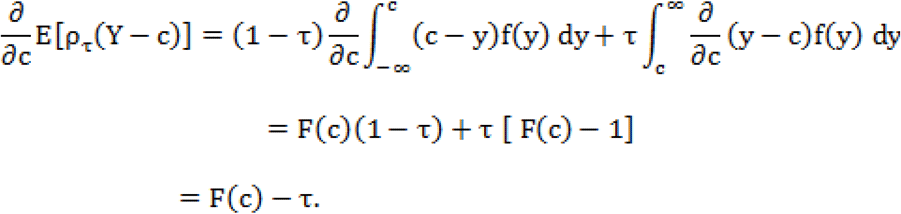

Now, setting a partial derivative equal to zero will lead to the results for the minimum:

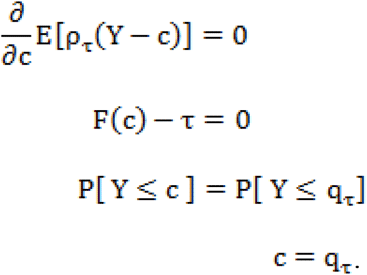

This solution is unique. Therefore, it can be written as c − F^−1^(τ). For τ = 0.5, we obtain the solution for the median. Now, for any 0 <τ< 1, it can be said that the τ^th^ sample quantile is the value of c that minimizes the sample expected loss function.

Let response variable y be associated with a set of explanatory variables x. Now, the concept of the unconditional mean can be extended to the conditional mean function estimation:

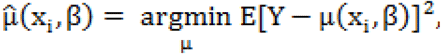

where, μ(x_i_,β) = E[ Y|X = x_i_] is termed as a conditional mean function. The conditional mean function of y given x is assumed linear. Therefore, a linear mean function can be written as, 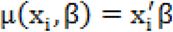. An ordinary least square solution can be defined as:

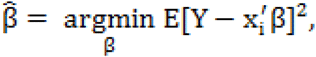

and now the τ^th^ sample quantile can be explained. Davino and his colleagues (2013) defined the τ^th^ conditional quantile function as:

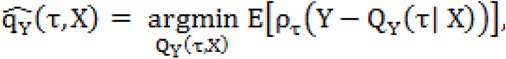

where, Q_Y_(τ| X) is defined as the conditional quantile function. In case of a linear quantile function, 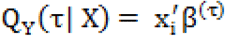. Here β^(τ)^ can be obtained by solving following equation:

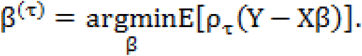

Suppose that random samples are drawn independently from a population and the resulting data structure is expressed as (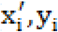); (i = 1,2,…, n). Here, y_i_ is a response variable (scalar) with a common CDF F F(y), and y_i_ is the p×1 vector of a design matrix X. Now, the τ^th^ linear conditional quantile can be modeled as:

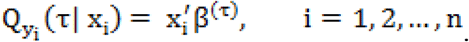

Here, 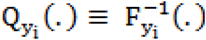 and β^(τ)^ is a column vector of length p with unknown fixed parameters. Now, the following minimization problem can be used to estimate β^(τ)^:

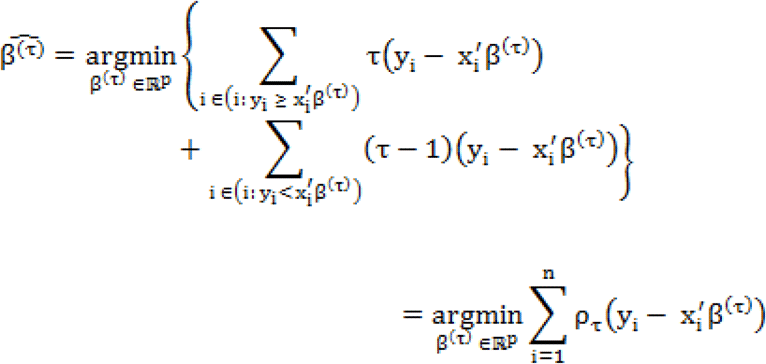

The above equation can be expressed in terms of a linear function subject to linear constraints. Thus, linear programming can be used to find β^(τ)^ (Koenker 1978). Here, parameter β^(τ)^ and its estimator 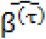 are both depend on τ ∊ (0,1).

## Results

The descriptive statistics of the mean precautionary score of the respondents of Dhaka obtained under different covariates are given in Table 1.

**Table 1.**
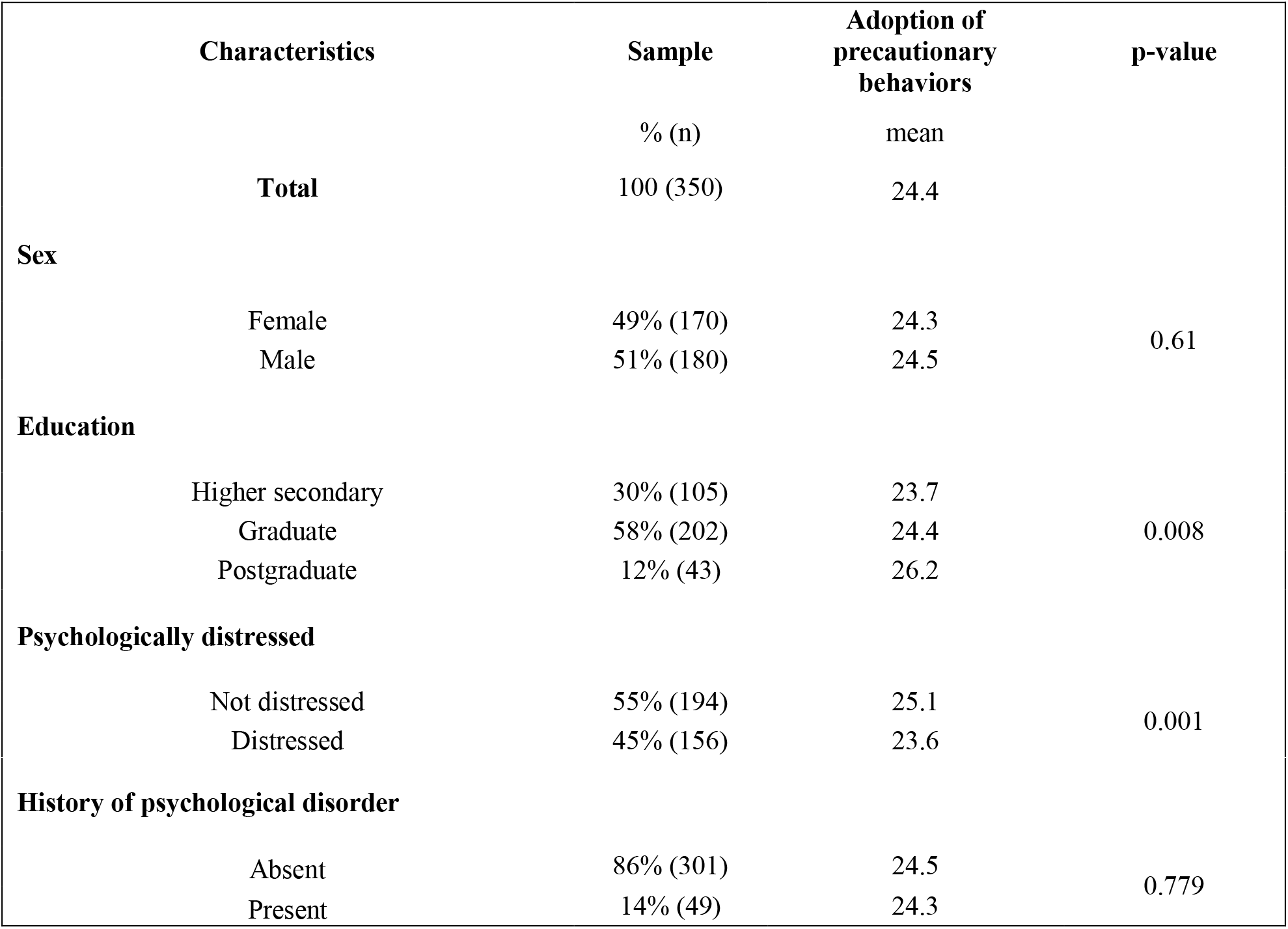
Sample distribution of Dhaka participants and mean adoption of precautionary behavior by demographic factors

To examine whether the mean precautionary score changed with the level of covariates, a two-sample t-test and analysis of variance (ANOVA) were conducted. In the resident of Dhaka, all covariates except sex and history of psychological disorder were found to have a significant association with precautionary score. The mean score varied significantly (p=0.008) with the level of education among the respondents of Dhaka. It was found that the mean precautionary score was highest (26.2) for the people who had completed their postgraduate education, whereas this mean score was 24.4 for those that completed the graduate level. The mean precautionary score was lowest (23.7) for those who had only a Higher Secondary Certificate (HSC) degree. Psychologically distressed people had lower mean (23.6) precautionary scores than those who were not distressed (mean=25.1). This result was found to be statistically significant at the 1% level of significance.

Table 2 shows that all covariates except educational level and history of psychological disorder were found to have a significant association with precautionary score among the resident of Tangail. The mean precautionary score of the young adults of Tangail differed significantly (p=0.007) by sex. Females (mean=25.3) proved to be more cautious than males (mean=23.8). Psychologically distressed participants had lower mean precautionary scores (23.7) than those who were not psychologically distressed (mean=25.1).

**Table 2.**
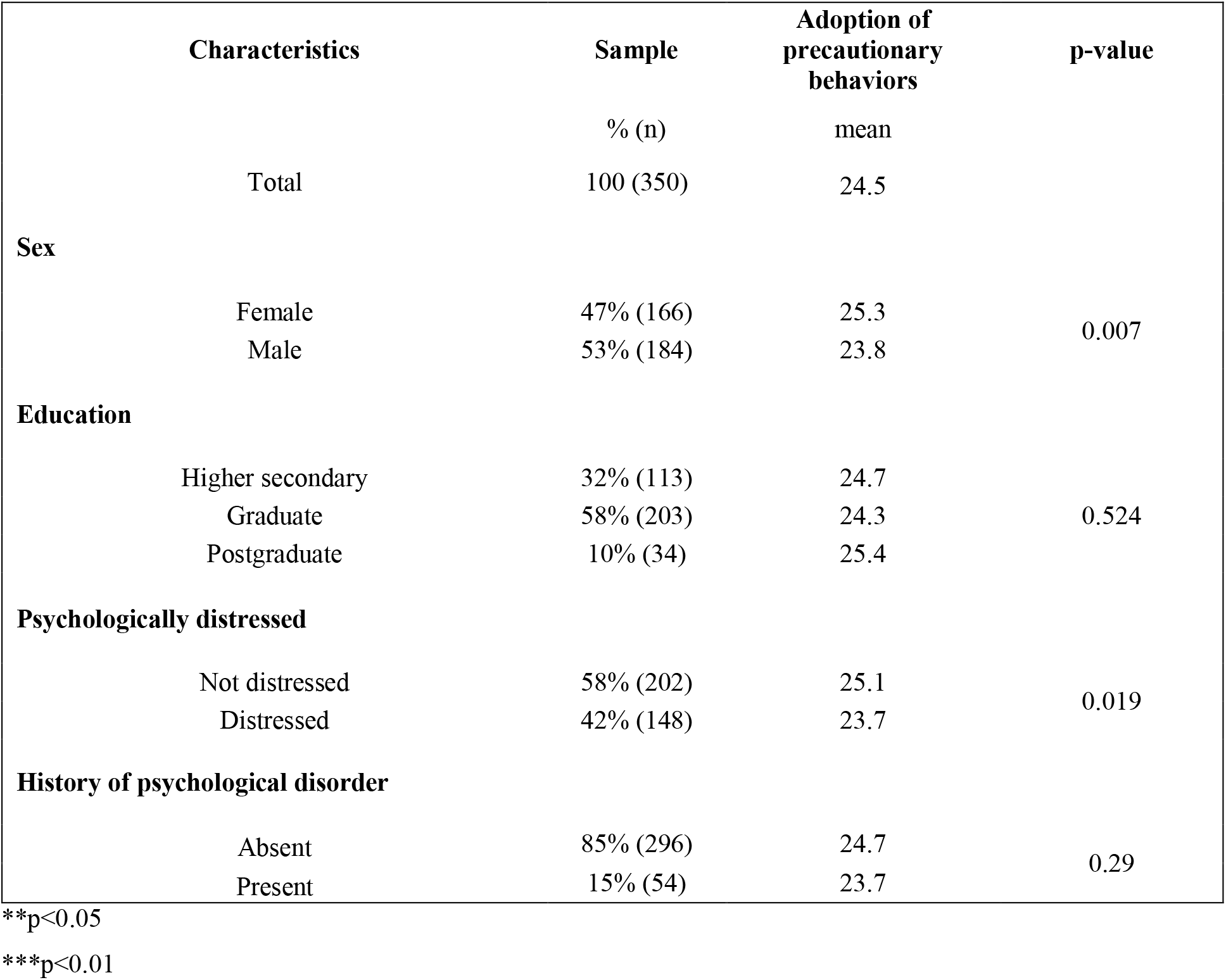
Sample distribution of the Tangail participants and mean adoption of precautionary behaviors by demographic factors

Table 3 shows that the mean precautionary score (24.4) of participants who were living Dhaka insignificantly differed from the mean precautionary score (24.5) of those who were living in Tangail.

**Table 3.** Differences in precautionary behaviors among people at the district level

| District | Sample<br>% (n) | Adoption of precautionary behaviors<br>mean | p-value |
| --- | --- | --- | --- |
| Dhaka | 50% (350) | 24.4 | **0.769** |
| Tangail | 50% (350) | 24.5 |  |
| **Total** | **100 (700)** | **24.48** |  |

Before fitting the regression model using the precautionary score as the dependent variable, it was essential to check whether the precautionary score data contained outliers. A box plot of the precautionary score is given in Fig. 1, and it is depicted from the figure that some observations of the precautionary score were detected as outliers for the sample of Dhaka, Tangail and the combined sample. Moreover, the skewness of the precautionary score was found to be -0.53, -0.82 and -0.71, indicating that the three distributions of the precautionary score were negatively skewed. Therefore, a linear regression model was not appropriate to examine the effects of covariates on the precautionary score for this scenario.

**Figure 1.**
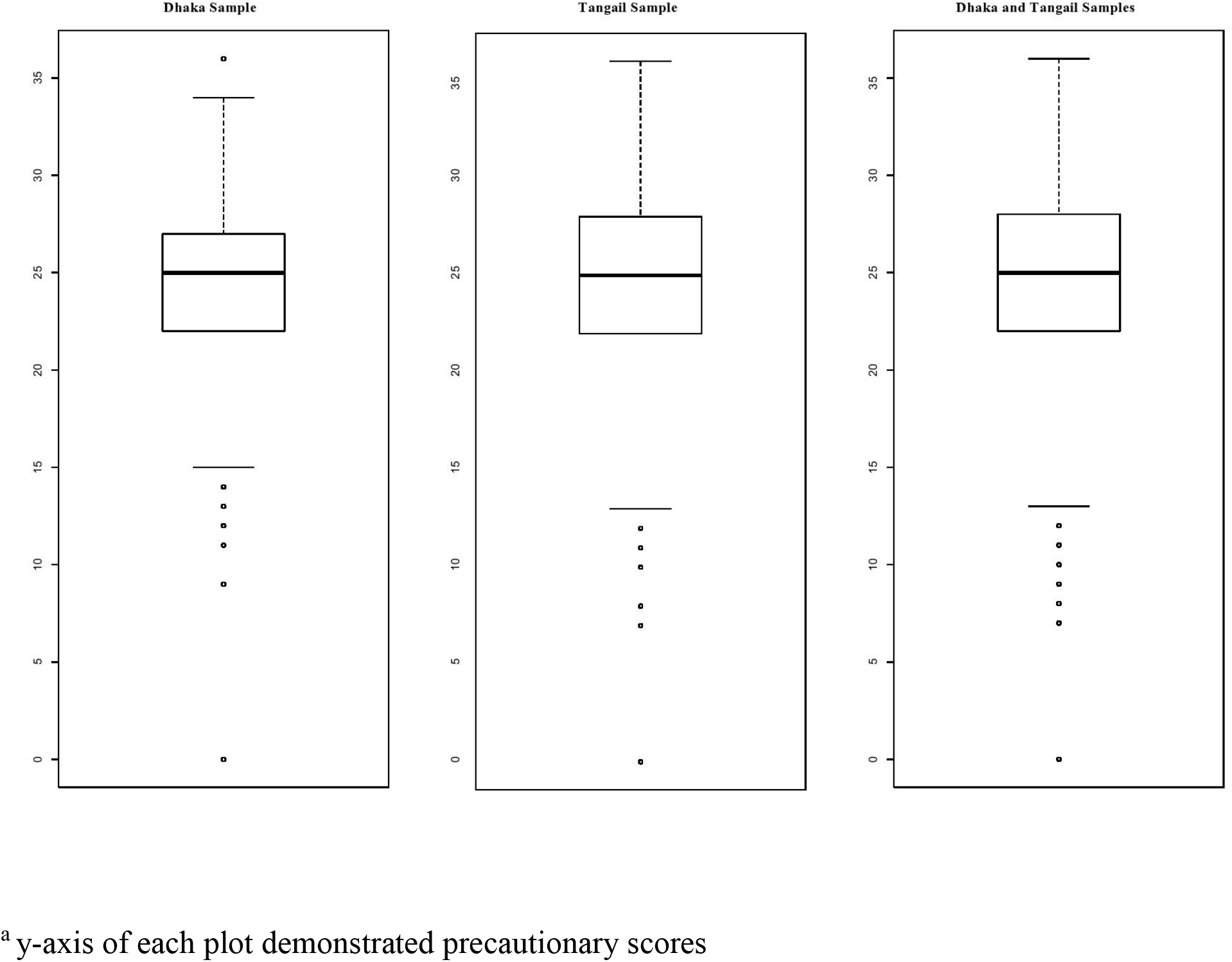
Boxplot for precautionary score

**Figure 2.**
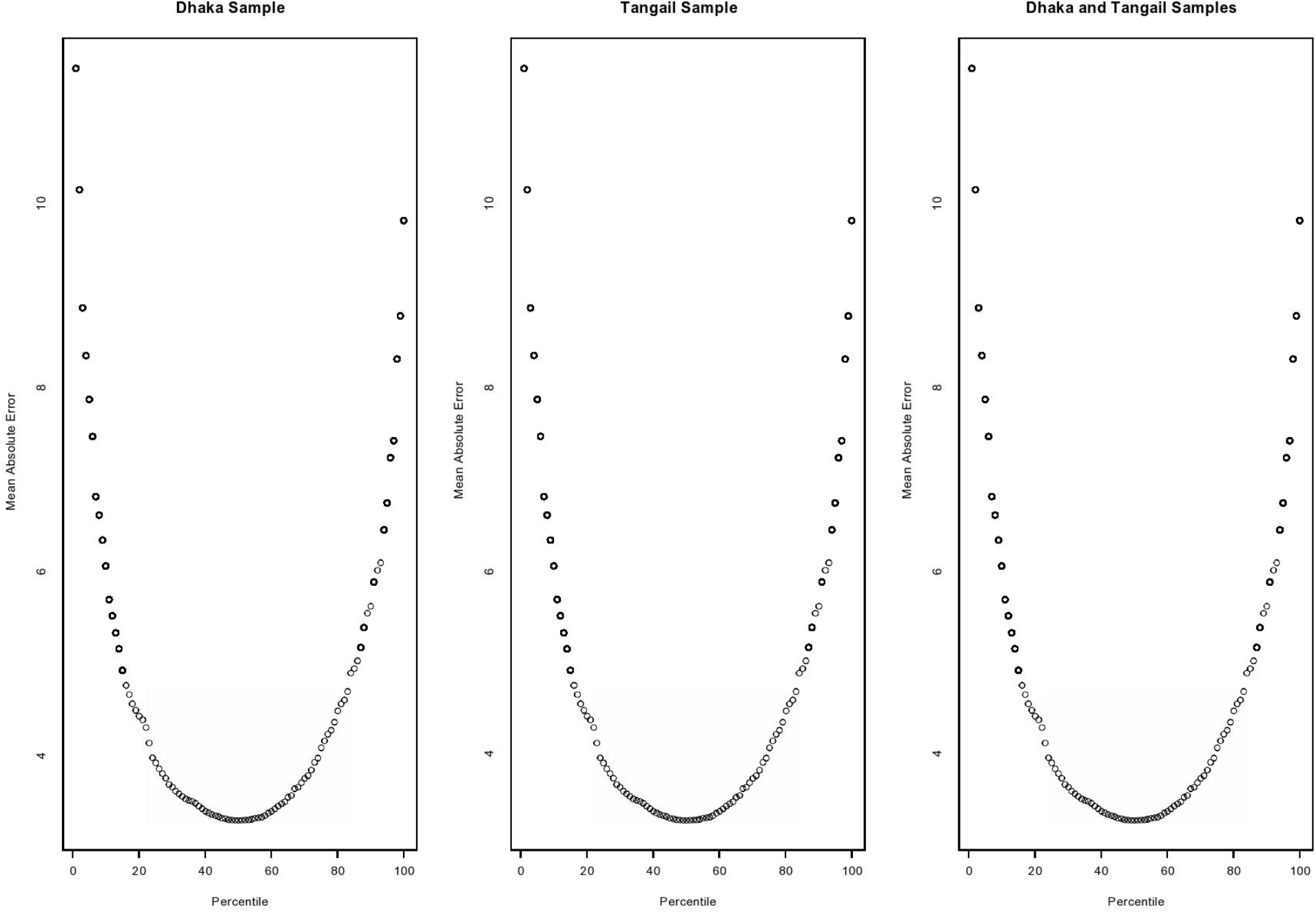
Mean absolute error (MAE) at different quantiles

Thus, to determine the adjusted contribution of the related variables on the precautionary score of people, a quantile regression model was fitted on the fiftieth quantile. From Fig. 2, it is evident that the value of the mean absolute error of our predicted values is lowest at the fiftieth quantile. Therefore, the median quantile regression model performs better in these scenarios to predict the precautionary score. Regression coefficients, standard errors, and p values are reported in Table 4. Model 1 is pertinent for the samples drawn from Dhaka; Model 2 is for the sample drawn from Tangail. Model 3 is the full sample quantile regression model combining samples from both districts in a single model.

**Table 4.** Quantile regression model for precautionary behavior adoption

| Variables | Model 1 (Dhaka sample) |  | Model 2 (Tangail sample) |  | Model 3 (Full sample) |  |
| --- | --- | --- | --- | --- | --- | --- |
|  | Coefficient (SE) | p-value | Coefficient (SE) | p-value | Coefficient (SE) | p-value |
| Intercept | 16.4 (2.02) | <0.001*** | 11.89 (1.95) | <0.001*** | 14.4 (1.22) | <0.001*** |
| Knowledge | 0.19 (0.15) | 0.23 | 0.12 (0.12) | 0.316 | 0.08 (0.08) | 0.333 |
| Attitude | 0.0 (0.24) | 1.0 | 0.08 (0.31) | 0.786 | 0.15 (0.18) | 0.392 |
| Self-control | 0.14 (0.02) | <0.001*** | 0.26 (0.03) | <0.001*** | 0.2 (0.02) | <0.001*** |
| Psychological distress | -0.9 (0.49) | 0.065* | -0.25 (0.5) | 0.61 | -0.32 (0.33) | 0.336 |
| **Sex (Ref: Male)** | - | - | - | - | - | - |
| Female | 0.04 (0.48) | 0.921 | -0.6 (0.48) | 0.212 | -0.06 (0.33) | 0.858 |
| **Education (Ref: Higher secondary)** | - | - | - | - | - | - |
| Graduate | 0.85 (0.55) | 0.119 | 0.28 (0.55) | 0.604 | 0.63 (0.4) | 0.117 |
| Postgraduate | 2.42 (0.57) | <0.001*** | 1.41 (0.96) | 0.142 | 2.3 (0.53) | <0.001*** |
| History of psychological disorder/chronic disease | -0.47 (0.66) | 0.474 | -0.26 (0.74) | 0.724 | -0.01 (0.41) | 0.978 |
| **District (Ref: Dhaka)** |  |  |  |  |  |  |
| Tangail | - | - | - | - | 0.02 (0.3) | 0.939 |

SE= standard error
\*p<0.1
\*\*\*p<0.01

As shown in Table 4, the self-control score was found to have a significant positive association with the precautionary score at the fiftieth quantile point in all three models [coefficient=0.14, 0.26, 0.2, respectively]. Postgraduate participants were estimated to be more precautious compared to their counterparts in model 1 and model 3 [coefficient=2.42, p<0.001; coefficient=2.30, p<0.001, respectively]. The precautionary score for psychologically distressed people was significantly lower [at the 10% level of significance; coefficient=-0.9, p=0.065] than the score for those who were not distressed in model 1. In summary, it can be concluded from the models that postgraduate participants with higher self-control and better mental health had developed better precautionary behaviors to prevent COVID-19.

## Discussion

The world right is now battling against an unforeseeable and unseen enemy because COVID-19 is highly transmissible even when the symptoms are absent (Bai et al. 2020; Li et al. 2020). The Secretary General of the United Nations has claimed this pandemic to be the most challenging event after the Second World War (PTI 2020). Circulating effective, sufficient, essential, updated, and accurate information regarding COVID-19 prevention and control has emerged as a major public health concern, and stopping this COVID-19 pandemic requires the utmost effort from every stakeholder along with frontline fighters and public health professionals (Srichan et al. 2020).

The United Nations Children’s Fund (UNICEF) has urged communities to take necessary protective actions and to support the control efforts so that this pandemic can be minimized (UNICEF 2020). In Beijing, the majority of the patients were young adults (Tian et al. 2020). South Korea experienced a low fatality rate amid a large outbreak of COVID-19 because the concentration of contamination was the highest among the young adults (Dowd et al. 2020).

Given this context, the primary goal of this research was to investigate which factors shape precautionary behaviors across different groups of young adults living in Dhaka and Tangail, Bangladesh. People with good mental health, i. e., less psychological distress, tended to adopt more preventive measures in the studied areas. The tendency of being less concerned about maintaining a healthy lifestyle among psychologically distressed persons is nothing new (Honda et al. 2005). Postgraduate participants in Dhaka reported the adoption of more precautions than the other participants. Female participants in Tangail with a better mental state were more cautious than males, which supports previous findings that men are more expected to exhibit risk-taking behavioral patterns (Duell et al. 2018; Pawlowski et al. 2008).

Among the variables selected for modeling, self-control was significantly associated with the adoption of precautionary behaviors in all three models. Education had a significant positive relationship with precautionary behavior in two models, and mental health became a significant predictor in one model. Knowledge, attitude, sex, and history of psychological disorder/chronic disease were not found to be effective predictors of precautionary behaviors.

Good knowledge about the high infectivity of COVID-19 makes people take a high level of precautions to prevent themselves from being contaminated (Zhong et al. 2020). Similarly, Li et al. (2020) found that knowledge of the participants regarding COVID-19 was positively related to their precautionary behavior adoption. Presumably, citizens of Dhaka have similarities in the magnitude of their access to information. Since knowledge is gained through information, invariance of knowledge among citizens made knowledge an insignificant predictor of precautionary behaviors. On the other hand, residents of Tangail might not have an equal opportunity to access the information or acquire knowledge from various sources. It was assumed that a person with more knowledge of COVID-19 may realize the importance of protective measures against this pandemic and do so accordingly in Tangail. Astoundingly, this relationship did not hold. Knowledge was not a factor that influences the adoption of precautionary behaviors in the studied areas.

Self-control refers to a person’s ability to change or dominate inner responses and to disrupt undesired behavioral tendencies (Tangney et al. 2004). Good self-control is obviously relevant to health outcomes, particularly in situations where the ability to restrain or initiate action is significantly related to the ability to handle difficulties arising from problematic situations (Li et al. 2020; Wills et al. 2008). This concept is reflected clearly in this study, as self-control was observed to be a significant predictor of precautionary behaviors in young adults in Dhaka and Tangail. Self-control positively induces a person to do what is right for him. Keeping a social distance, using protective equipment appropriately, maintaining cough etiquette, and quarantining oneself even if minimal symptoms are present are similar to fighting a war during this pandemic era. A proper precautionary step taken by one person can save the lives of many. A person with better self-control is, therefore, a better fighter against COVID-19 since he is more likely to adopt precautionary behaviors. In many cases, a higher level of education was associated with a higher level of precautionary behavior. A postgraduate person seemingly possesses better knowledge and understanding of what to do in this sort of never-seen situation, which is why it was well anticipated that a postgraduate resident of Bangladesh would be more precautious than others.

It is evident that the perceived personal risk of infection and health effects can predict individuals’ engagement in protective behavior (Bish and Michie 2010). A very recent study identified a subgroup that was less engaged in protective behavior. This cohort consisted of people with disengagement in information-seeking who thought the situation was less risky than others thought; the members of this cohort thought they would not be affected by COVID-19 (Wise et al. 2020). These characteristics indicate that less informed individuals misperceive the intensity of the situation. Moreover, these individuals either do not realize the danger of the situation or mistakenly consider themselves immunized against the danger. Maintaining a proper attitude toward emergency situations is therefore very necessary. An optimistic attitude toward disease can result in the success of control measures such as traffic limits and location lockdowns (Zhong et al. 2020). During the SARS outbreak, optimistic thinkers more often avoided crowded or public places but portrayed poor adoption of health behaviors, such as less use of disinfectants and washing their hands infrequently. On the other hand, those who were more concerned about SARS reported better adoption of health behaviors and less avoidance (Lee-Baggley et al. 2004). Surprisingly, attitude did not appear to be a significant predictor of precautionary behavior in any of the samples in this study.

Participants from Dhaka and Tangail, in general, showed moderate precautionary behavior during the rising period of COVID-19 in Bangladesh.

Raising awareness by penetrating actual, effective, and timely information regarding COVID-19 to the leaders among its citizens can be one of the short-term focuses of the Bangladeshi government. The circulation of false information, rumors, and unauthentic news from different social networking sites and online news portals obviously misguides people and, in effect, promotes the loss of self-control. These factors can result in unanticipated fatal outcomes. Publicizing scientific information about COVID-19 as thoroughly as possible can enhance the development of the public’s healthy behaviors (Zhong et al. 2020). Appropriate filtration of information, exclusion of rumors, and meticulous penetration and filtration of information into the public’s cognition are all needed in Bangladesh before lifting the lockdown up. As containing better knowledge and a proper attitude cannot ensure the adoption of more precautionary behaviors among the people, the government’s strong intervention to keep people at home and distant from each other while continuing the lockdown for several more days may be the immediate solution. At the same time, the economic burden on the lower-income residents should be addressed so that people do not have to suffer starvation due to the shutdown of economic activities.

## Limitations

Even though the findings of this early study can bear several implications, both theoretically and empirically, it could not overcome some limitations. The study covered only educated young adult participants and could not incorporate the responses from elderly people living in remote areas. Rural people and older people are more likely to have inappropriate precautionary behaviors, and they need special research efforts. Analyses of a more representative sample drawn from all over Bangladesh can show more vigorous findings.

## Data Availability

Data is submitted to a data repository.

https://data.mendeley.com/datasets/rsccrhfsz7/2

## Compliance with ethical standards

### Funding

No funding was received for this study.

## Authors’ contribution

Asif Imtiaz contributed to the study conception and design. Material preparation, data collection and analysis were performed by Asif Imtiaz, and Noor Muhammad Khan. The first draft of the manuscript was written by Asif Imtiaz and Noor Muhammad Khan. Md. Akram Hossain approved the final version to be published. All authors read and approved the final manuscript.

## Ethical Approval

The study was approved by the research ethics committee of Center for Project Management and Information Systems (PMIS), University of Dhaka. The procedures used in this study adhere to the tenets of the Declaration of Helsinki.

